# Effects of mobility and multi-seeding on the propagation of the COVID-19 in Spain

**DOI:** 10.1101/2020.05.09.20096339

**Authors:** Mattia Mazzoli, David Mateo, Alberto Hernando, Sandro Meloni, José J. Ramasco

## Abstract

Assessing the impact of mobility on epidemic spreading is of crucial importance for understanding the effect of policies like mass quarantines and selective re-openings. High mobility between areas contribute to the importation of cases, affecting the spread of the disease. While many factors influence local incidence and making it more or less homogeneous with respect to other areas, the importance of multi-seeding has often been overlooked. Multi-seeding occurs when several independent (non-clustered) infected individuals arrive at a susceptible population. This can give rise to autonomous outbreaks that impact separate areas of the contact (social) network. Such mechanism has the potential to boost local incidence and size, making control and tracing measures less effective. In Spain, the high heterogeneity in incidence between similar areas despite the uniform mobility control measures taken suggests that multi-seeding could have played an important role in shaping the spreading of the disease. In this work, we focus on the spreading of SARS-CoV-2 among the 52 Spanish provinces, showing that local peaks of incidence and mortality strongly correlate with mobility occurred in the early-stage weeks from and to Madrid, the main mobility hub and where the initial local outbreak unfolded. These results clarify the higher order effects that mobility can have on the evolution of an epidemic and highlight the relevance of its control.

## INTRODUCTION

The COVID-19 epidemic, firstly detected in the Hubei province in China, reached the status of pandemic on March 11, 2020 [1] and currently involves at least 187 countries around the entire world with almost 3.5 million cases and more than 240, 000 deaths as of the beginning of May 2020 [2].

In Europe, SARS-CoV-2 severely hit Italy, the first country to report local transmission, in mid-February [3] and, by the end of the month, other European countries like Spain were already experiencing sustained local outbreaks [3]. Madrid was the first region to confirm large numbers of local contagions and, on March 6, 2020, the city already reached 100 confirmed cases. This led to the closure of public buildings and different activities, like, for instance, universities, which brought the lateral effect of getting people who reside in the city for work or study to leave and go back to their province of origin. As most major capitals in the world, Madrid attracts workers and students from all the country, a large fraction of daily commuters from neighboring provinces and weekly travelers from all provinces. Among these for example Soria, one of the most affected provinces by COVID-19 per capita, which counts with a scarce population and density but a large number of weekly commuters with the capital. On March 14, given the fast spread of the virus in the country, the Spanish Government declared the Alarm State (Estado de Alarma) [4]. Such State allows the Government to impose restrictions on economic activities and citizens mobility. The restrictions included a reduction of public transport services like buses, metros and trains around the 50% of the usual capacity, the closure of buildings and activities related to the public sector and the limitation of individual mobility in the whole country. People could leave home only for essential needs as grocery shopping, visiting the doctor and pharmacies, or going to work. Mobility between provinces was limited to working reasons. This compressed the closing of many economic activities, especially those related to the service sector, letting only some exceptions like logistics and transport, supermarkets, pharmacies, health-related centers, construction, agriculture and industry to keep operating. On March 29, given the growth in the number of new cases, the Government declared the closing of all non-essential activities. After April 13, 2020, the restriction measures have started to be progressively lifted but we will center on the initial stages of the spreading.

In this work, combining detailed human mobility data with epidemiological reports, we study the relationship between inter-city mobility flows and COVID-19 incidence. Our working hypothesis is that large mobility flows lead to multiple seeding events resulting in several local outbreaks and, finally, to a higher incidence. Although the role of human mobility in shaping epidemic dynamics has been extensively considered [5-14], even recently for COVID-19 [15-21], the effect of multiple seeding due to mobility has received sensibly less attention; with only few theoretical [22, 23] and applied exceptions [24-27]. To test this assumption, we focus on the change in mobility among the province of Madrid - where the first sustained outbreak was recorded- and the other provinces in Spain the week before the onset of the local outbreaks. The same analysis is then performed on the mortality instead of the incidence, attesting the robustness of our approach. Our results confirm that trips between Madrid and other provinces occurred one week before the onset explain the 67% of the peak incidence for each area. Moreover, we find that a model that takes into account trips, population size and onset time accounts for the 77% of the SARS-CoV-2 incidence and mortality during the peak in each area of the country.

## DATA AND METHODS

We take as basic geographical units Spanish provinces (see Fig. 1), which are 50 territorial divisions and 2 autonomous cities in the Northern coast of Africa (Ceuta and Melilla). They correspond to the NUTS 3 statistical areas according to the European nomenclature.

**FIG. 1.**
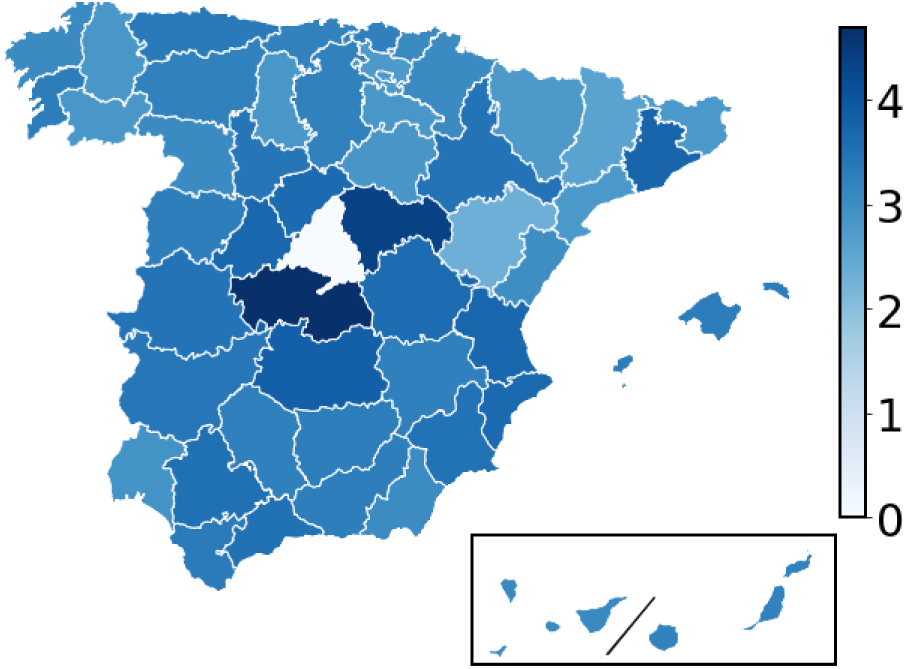
Division of Spain in provinces. The color code corresponds to the decimal logarithm of the number of residents in each province with visits to Madrid (white region in the center) on March 4.

The characterization of inter-province mobility has been inferred from anonymized records of telecommunication network activity by Kido Dynamics SA (www.kidodynamics.com). The dataset contains approximately 13 million devices with unique daily mobility patterns, each being composed of a series of trips and visits. A *trip* is defined as a sequence of records per device that are only compatible with displacements of at least 5 *km/h* in the past 1 hour window. Trips are aggregated by origin (destination) province according to the location of the antennas where the first (last) records occurred. Complementary to trips, a *stay* is defined as any sequence of consecutive records that do not constitute trips. Stays are aggregated by province according to the location of the antennas where all their records occurred. Each device is assigned a residence province according to the most common location of its stays outside office hours. All the stays outside a device’s residence area are then labeled as “visits”. Figure 1 shows, for instance, the number of residents from the different provinces with stays in Madrid. Mobility patterns in Spain display significantly different behaviors during the weekend and weekdays. Thus, we classify the visits of a given device to a given province as “weekender” if more than half of the device’s visits to that province during the period of study occurred during the weekend.

Sequence of labeled trips and visits are then aggregated daily into a total number of trips per province of residence and province of origin/destination, and total number of visits per province and province of residence, both for weekenders and for non-weekenders. Finally, inspired by differential privacy methods [28] and to avoid any potential disclosure of personally-identifiable mobility patterns, a small unbiased Laplacian noise with scale parameter of 5 (i.e. variance of 50) is added to all aggregated values. After adding the noise, any values below 10 are discarded from the sample as additional preventive measure. Such aggregated trip and stays information are the basis for the mobility analysis presented in this work.

Regarding the number of cases, the incidence and the mortality at the peak, the data has been collected from official health services releases where available. The list of links is at Ref. [29].

## RESULTS

We define a group of key features of the spreading directly from the epidemic curves of incidence and mortality per day from the beginning of the epidemic to April 13, 2020, which is when testing policies changed, rising sensibly the coverage of registered cases. A first word of warning is required: the data of both curves are noisy due to changes in the testing rhythms. It is easy, for instance, to detect the effect of weekends with a clear slowing down of testing and reporting. A process of smoothing is, hence, applied to obtain more reliable estimates on the trends of the epidemic. To do this, we take a running average of three days assigning the value to the central point.

Once the curves have been smoothed-out (see Fig. 7 below), we record the magnitude of the peak, the times between the local onsets (when the incidence is larger than 7/100, 000) and the respective peaks in every province (∆τ), and the time between the local onset and March 6 (∆*t_M_*), which is when Madrid reached 100 confirmed cases. Four provinces (Almeriia, Cadiz, Huelva, Las Palmas and Sevilla) have few detected cases and do not reach the established onset, thus we have excluded them from the first analysis on times. The list of onset and peak times for the incidence can be found in Table I.

**TABLE I.** Table of epidemic features for the 52 provinces of Spain, when available.

| Province | Incidence onset | Incidence peak time | Mortality onset | Mortality peak time |
| --- | --- | --- | --- | --- |
| Álava | 2020-03-07 | 2020-03-25 | 2020-03-16 | 2020-03-31 |
| Albacete | 2020-03-17 | 2020-03-29 | 2020-03-17 | 2020-04-05 |
| Alicante | 2020-03-24 | 2020-03-29 | 2020-03-21 | 2020-04-03 |
| Almería | nan | 2020-03-28 | 2020-03-22 | 2020-04-10 |
| Ávila | 2020-03-18 | 2020-03-27 | 2020-03-17 | 2020-03-26 |
| Badajoz | 2020-03-25 | 2020-03-27 | 2020-03-24 | 2020-04-09 |
| Baleares | 2020-03-22 | 2020-03-26 | 2020-03-21 | 2020-04-01 |
| Barcelona | 2020-03-15 | 2020-03-25 | 2020-03-12 | 2020-03-31 |
| Burgos | 2020-03-19 | 2020-03-24 | 2020-03-16 | 2020-03-31 |
| Cáceres | 2020-03-19 | 2020-03-27 | 2020-03-14 | 2020-04-01 |
| Cádiz | nan | 2020-04-01 | 2020-03-27 | 2020-04-08 |
| Castellón | 2020-03-23 | 2020-03-26 | 2020-03-21 | 2020-04-08 |
| Ciudad Real | 2020-03-17 | 2020-04-04 | 2020-03-17 | 2020-04-09 |
| Córdoba | 2020-03-28 | 2020-04-04 | 2020-03-19 | 2020-03-31 |
| A Coruña | 2020-03-22 | 2020-04-01 | 2020-03-22 | 2020-04-01 |
| Cuenca | 2020-03-19 | 2020-04-08 | 2020-03-17 | 2020-03-27 |
| Girona | 2020-03-18 | 2020-03-25 | 2020-03-17 | 2020-04-03 |
| Granada | 2020-03-23 | 2020-03-30 | 2020-03-16 | 2020-03-28 |
| Guadalajara | 2020-03-16 | 2020-03-31 | 2020-03-21 | 2020-03-25 |
| Guipúzcoa | 2020-03-22 | 2020-03-25 | 2020-03-20 | 2020-04-10 |
| Huelva | nan | 2020-04-02 | 2020-03-27 | 2020-04-06 |
| Huesca | 2020-03-23 | 2020-04-01 | 2020-03-21 | 2020-04-09 |
| Jaén | 2020-03-23 | 2020-03-30 | 2020-03-13 | 2020-04-08 |
| León | 2020-03-22 | 2020-03-28 | 2020-03-16 | 2020-04-03 |
| Lleida | 2020-03-18 | 2020-03-25 | 2020-03-16 | 2020-03-26 |
| La Rioja | 2020-03-07 | 2020-03-27 | 2020-03-11 | 2020-04-06 |
| Lugo | 2020-03-25 | 2020-04-01 | 2020-03-23 | 2020-04-01 |
| Madrid | 2020-03-09 | 2020-03-25 | 2020-03-09 | 2020-03-27 |
| Málaga | 2020-03-24 | 2020-03-24 | 2020-03-14 | 2020-04-10 |
| Murcia | 2020-03-25 | 2020-03-25 | 2020-03-24 | 2020-04-05 |
| Navarra | 2020-03-14 | 2020-03-26 | 2020-03-15 | 2020-03-30 |
| Ourense | 2020-03-22 | 2020-04-01 | 2020-03-22 | 2020-04-01 |
| Asturias | 2020-03-18 | 2020-03-19 | 2020-03-19 | 2020-04-10 |
| Palencia | 2020-03-20 | 2020-03-27 | 2020-03-24 | 2020-04-04 |
| Las Palmas | nan | 2020-03-28 | 2020-03-20 | 2020-03-21 |
| Pontevedra | 2020-03-21 | 2020-04-01 | 2020-03-26 | 2020-03-31 |
| Salamanca | 2020-03-17 | 2020-03-28 | 2020-03-16 | 2020-03-28 |
| Santa Cruz de Tenerife | 2020-03-24 | 2020-03-26 | 2020-03-20 | 2020-03-21 |
| Cantabria | 2020-03-19 | 2020-03-26 | 2020-03-20 | 2020-03-31 |
| Segovia | 2020-03-17 | 2020-04-07 | 2020-03-16 | 2020-03-30 |
| Sevilla | nan | 2020-03-28 | 2020-03-22 | 2020-04-07 |
| Soria | 2020-03-17 | 2020-03-25 | 2020-03-16 | 2020-03-30 |
| Tarragona | 2020-03-24 | 2020-03-31 | 2020-03-21 | 2020-04-08 |
| Teruel | 2020-03-23 | 2020-03-26 | 2020-03-16 | 2020-04-01 |
| Toledo | 2020-03-17 | 2020-03-26 | 2020-03-17 | 2020-04-09 |
| Valencia | 2020-03-23 | 2020-03-24 | 2020-03-21 | 2020-04-02 |
| Valladolid | 2020-03-19 | 2020-04-10 | 2020-03-18 | 2020-04-01 |
| Vizcaya | 2020-03-19 | 2020-03-25 | 2020-03-17 | 2020-04-08 |
| Zamora | 2020-03-22 | 2020-04-03 | 2020-03-17 | 2020-04-03 |
| Zaragoza | 2020-03-23 | 2020-03-26 | 2020-03-10 | 2020-04-01 |
| Ceuta | 2020-03-30 | 2020-03-30 | 2020-03-26 | 2020-04-05 |
| Melilla | 2020-04-03 | 2020-04-03 | 2020-03-24 | 2020-03-24 |

On the mobility side, we define the total trips divided by the province population (intended as the sum of inflow and outflow) between Madrid and each province one week before the local onset. For example, if the onset in province A occurred on March 15, 2020, the trips counted are those registered between March 1 and 7. We checked the correlations between the local epidemic metrics and the mobility in different time ranges but one week before the local onset produces the best results. For the sake of completeness, we have also considered the mobility of a single day seven days before the local onset.

We start our analyses by confirming some correlations of the incidence that one could a priori expect. As can be seen in Fig. 2a, the height of the incidence peak is connected to the time between the local onset and peak in every province. This is natural, but it testifies that the outbreaks behave similarly across provinces once local contagion ignites. An analogous, even clearer relation, can be observed between the maximum incidence and the delay of the local onset from the arrival of Madrid at 100 cases (Fig. 2b). As can be observed in Figs. 2c and d, the time of the local onset is highly related to the total trips from/to Madrid the week before, showing that mobility was the main driver of the epidemic expansion. The main outliers here are La Rioja and Alava, which started the onset right after Madrid without having such high level of trips per capita with the capital. This is due to a super-spreading event registered in a wedding that strongly boosted local contagion in both neighboring provinces almost at the same time that the epidemic started to ramp up in Madrid [30]. Finally and more importantly for the hypothesis of the relevance of multi-seeding, Figs. 2e and f show the connection between the height of the incidence peak and the trips from/to Madrid weighted by the local population of the province with a high level of correlation. There are some outliers as the provinces neighboring Madrid (Avila, Cuenca, Guadalajara and Toledo), which fall within the functional metropolitan area of the capital and thus behave similarly to Madrid itself. This explains the flattening of the trends on the right side of the plots.

**FIG. 2.**
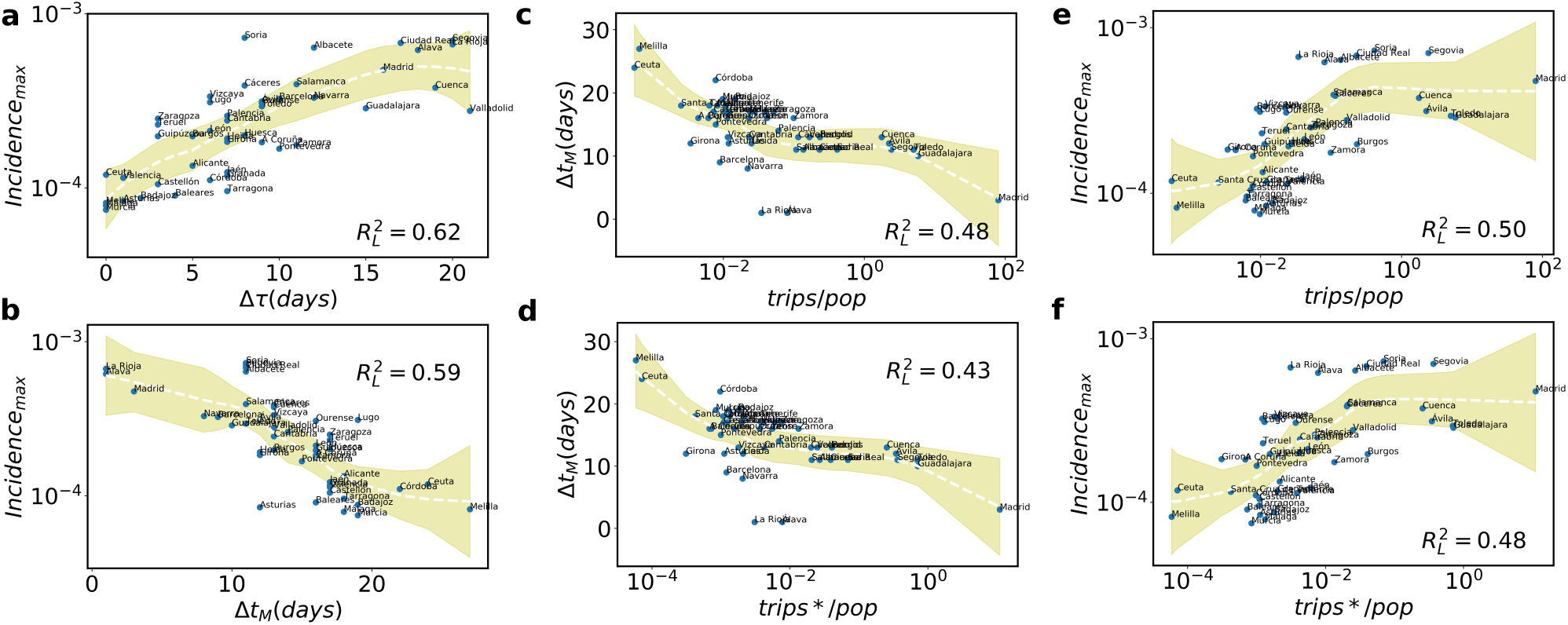
Correlations of the incidence. **a** Incidence at the peak as a function of the time between local onset and peak (∆τ); **b** Relation between the incidence at the peak and the time difference between the local onset and the case number 100 detected in Madrid (∆*t_M_*); c Delay in the onset ∆*t_M_* as a function of the trips from/to Madrid one week before the local onset; d The same but with the trips of a single day seven days before the local onset; e Peak of incidence versus the trips divided by the local population to/from Madrid one week before the local onset. f The same (peak of incidence vs mobility) but instead of taking a full week only seven days before the local onset. In all cases, the curves with the fits are obtained using the non parametric LOESS method, as also the values of 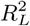. The shaded areas correspond to the 95% confidence interval.

The picture is maintained if one focuses on the mortality instead of on the incidence. The local onset for the mortality is established when every province reaches 1/1000 000 (see Fig. 8 and Table I below). Differently from before, the height of the peak of mortality is not so strongly connected to the time between the local mortality onset and the peak (see Fig. 3a). The peak of mortality correlates better with the time difference between the local onset and March 6 (Fig. 3b). The mobility for the analysis is the same as that taken for the incidence, one week before or 7 days before the onset of the incidence in every province. The results of plotting the time difference between the local mortality onset and March 6 are shown in Fig. 3c (for the one week mobility) and in Fig. 3d for a single day mobility. In general, the higher the mobility between the province and Madrid, the faster casualties are observed. This relation is valid for all provinces except for those with a strong overlap with the metropolitan area of Madrid, which once again are outliers in these plots. As shown in Figs. 3e and f, the mobility the week before the local incidence onset from/to Madrid in trips per capita of the province correlates with the peak in mortality. This further contributes to the hypothesis that the arrival of multiple-seeds can increase the local size of the epidemic and, consequently, the mortality.

**FIG. 3.**
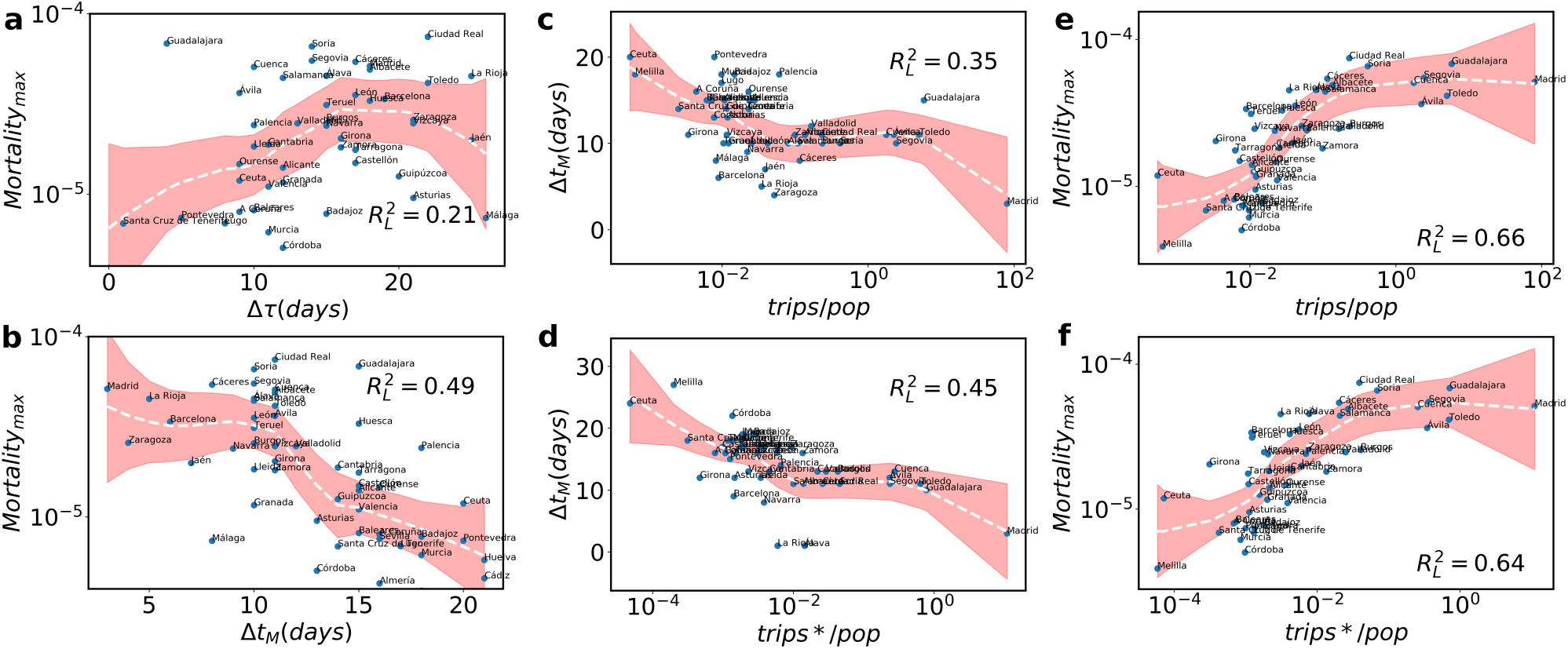
Correlations for mortality. **a** Mortality at the peak as a function of the time between the local mortality onset (> 1/1 000 000) and the peak. **b** Mortality peak versus the time between the local onset and March 6. **c** Time between the local mortality onset and March 6 versus the trips from/to Madrid one week before the onset. **d** The same but with the trips of one single day seven days before the local onset. **e** Relation between the height of the mortality peak and the trips from/to Madrid divided by the local province population. **f** Mortality peak versus the trips per capita from/to Madrid seven days before the onset. As before, the fits and the value of 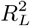 are obtained with LOESS. The shaded areas correspond to the 95% confidence interval.

We have analyzed so far total trips from/to Madrid but one person can be responsible for more than one of these trips. The status of each single individual against the pandemic is an intrinsic variable and, therefore, to improve the results it is necessary to consider the stays instead of the trips. This allows us to analyze the number of stays of residents in Madrid in the different provinces and, the other way around, the number of residents of those provinces that visited Madrid and came back home. In all the cases and as above, we will take as basis the mobility one week before the onsets in incidence in the different provinces. Out of different types of mobility that occur between provinces and the capital, we separate it into two classes: the visits of weekenders to the provinces and visits of weekday travelers. In Fig. 4, one can observe the correlation between the height of the local incidence and mortality and the stays per capita of weekenders and weekday travelers in the different provinces one week before the corresponding onsets. Neighboring provinces overlapping with Madrid metro area have higher commuting levels (weekday visits) to the capital with respect to others. This is understandable given that for them the trips are internal mobility. Such effect brings the saturation in the curves of Fig. 4b and c. On the other side, weekender stays from/at Madrid are not affected by distance as much as for weekday visitors, hence the saturation for neighboring provinces disappears. By this analysis, we can understand that those provinces where a higher relative peak of contagions and deaths occurred exhibit higher visits to/from the capital per capita the week before the onsets and, therefore, a higher inflow of seeds. Note that we do not find differences in considering residents of Madrid visiting the provinces or province residents traveling to Madrid, the effect is the same.

**FIG. 4.**
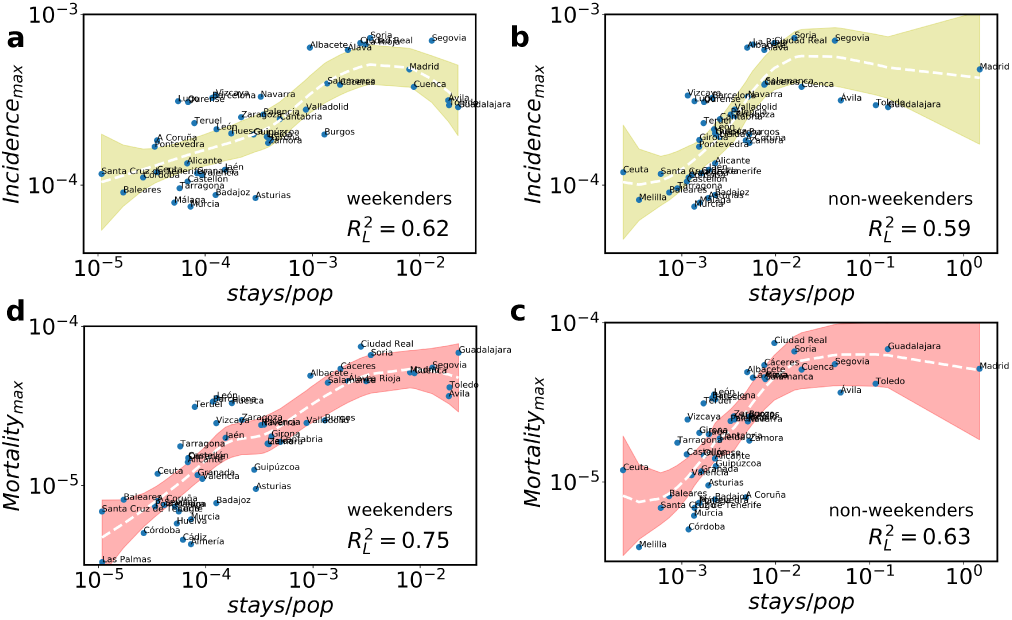
Stay analysis. The height of the local incidence peak as a function of the visits per capita of a weekenders and b weekdays travelers from/to Madrid one week before the incidence onset. In c (weekenders) and d (weekday travelers), the same analysis but for the peak of mortality and with the stays of one week before of the incidence onset. The curves and 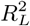 correspond to the LOESS non-parametric fit, shaded ares are the 95% confidence intervals.

Besides the number of stays per capita, we can analyze as well the duration of the visits to check whether longer stays correlate better with the magnitude of the peaks. However, changes in the correlation are so small that the information given by the duration of stays is negligible. It is thus not so relevant to separate the visitors either by origin or stay duration.

To further understand the relevant variables contributing to the heights of the local peaks of incidence and mortality, we perform a multivariate analysis including five variables: logarithm of total stays per capita one week before the incidence onset, density of population, distance from Madrid, population of every province and onset times. Note that the onset time is an intrinsic epidemic variable and as such it can depend on the others (e.g., mobility). For those provinces that did not reach the established onset, here we consider the peak day as a proxy for the onset. As shown in Figure 5, mobility per capita alone explains the 67% (incidence) and 68% (mortality) of the peak heights variance. By adding all the variables for each province, the explained variances increase to 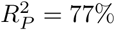 and 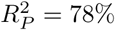, respectively.

**FIG. 5.**
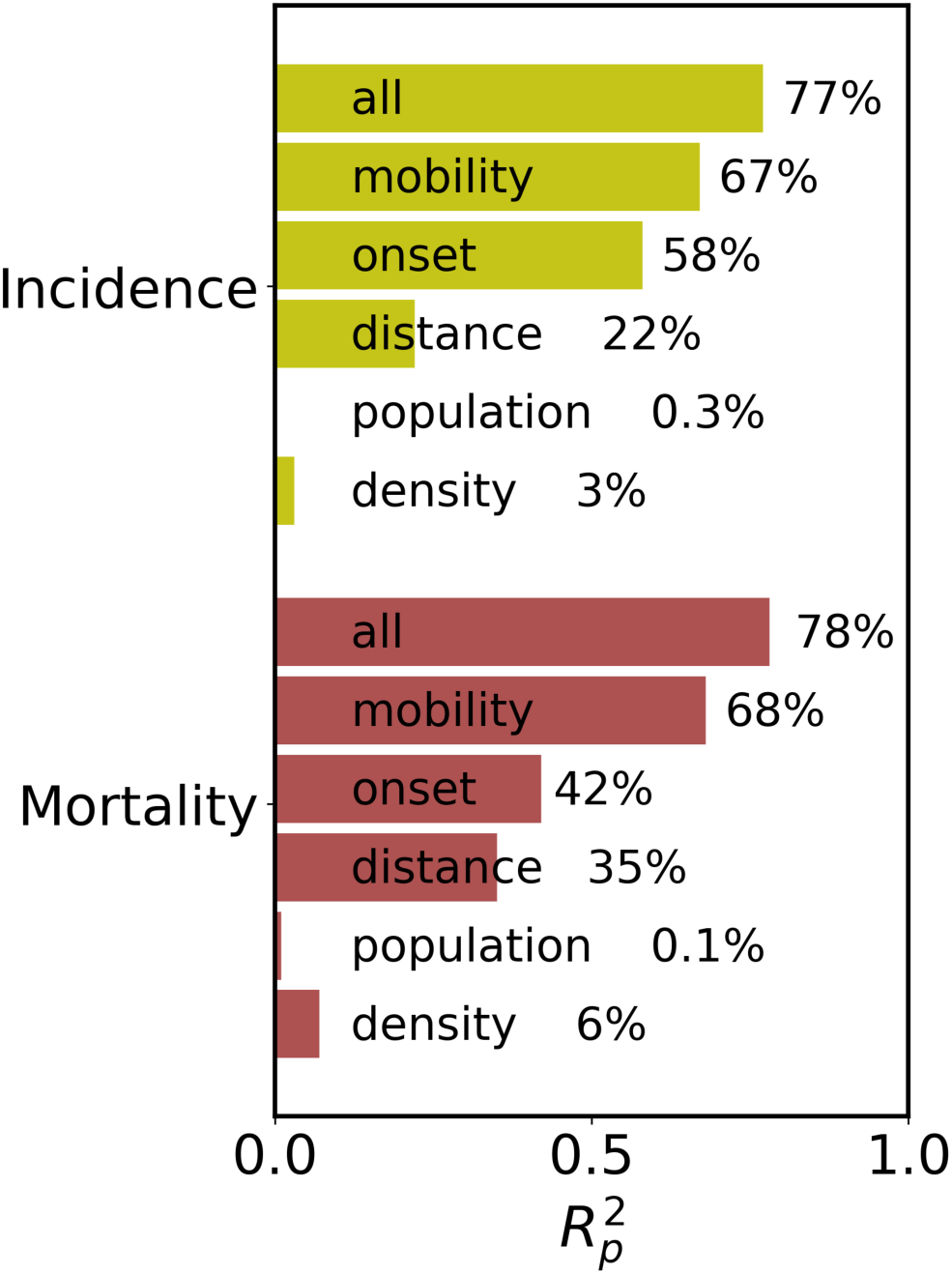
Multivariate analysis. Bar chart with the multivariate correlation results in 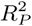 for the incidence and mortality peak heights in each province as a function of the mobility per capita and onset times. In every case, the upper bar corresponds to the simple correlation with mobility, below the multivariate correlation with onset time, onset time plus population and finally all the three variables together. In this analysis, 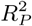 refers to R-squared of the linear Pearson correlation.

By using the multivariate fitted parameters, we can reconstruct the estimated incidence and mortality peaks for each province and compare the statistical model predictions and the actual values. As illustrated in Figure 6, both statistical models (for incidence and mortality) give good accuracy in the estimation of the real peaks for each province.

**FIG. 6.**
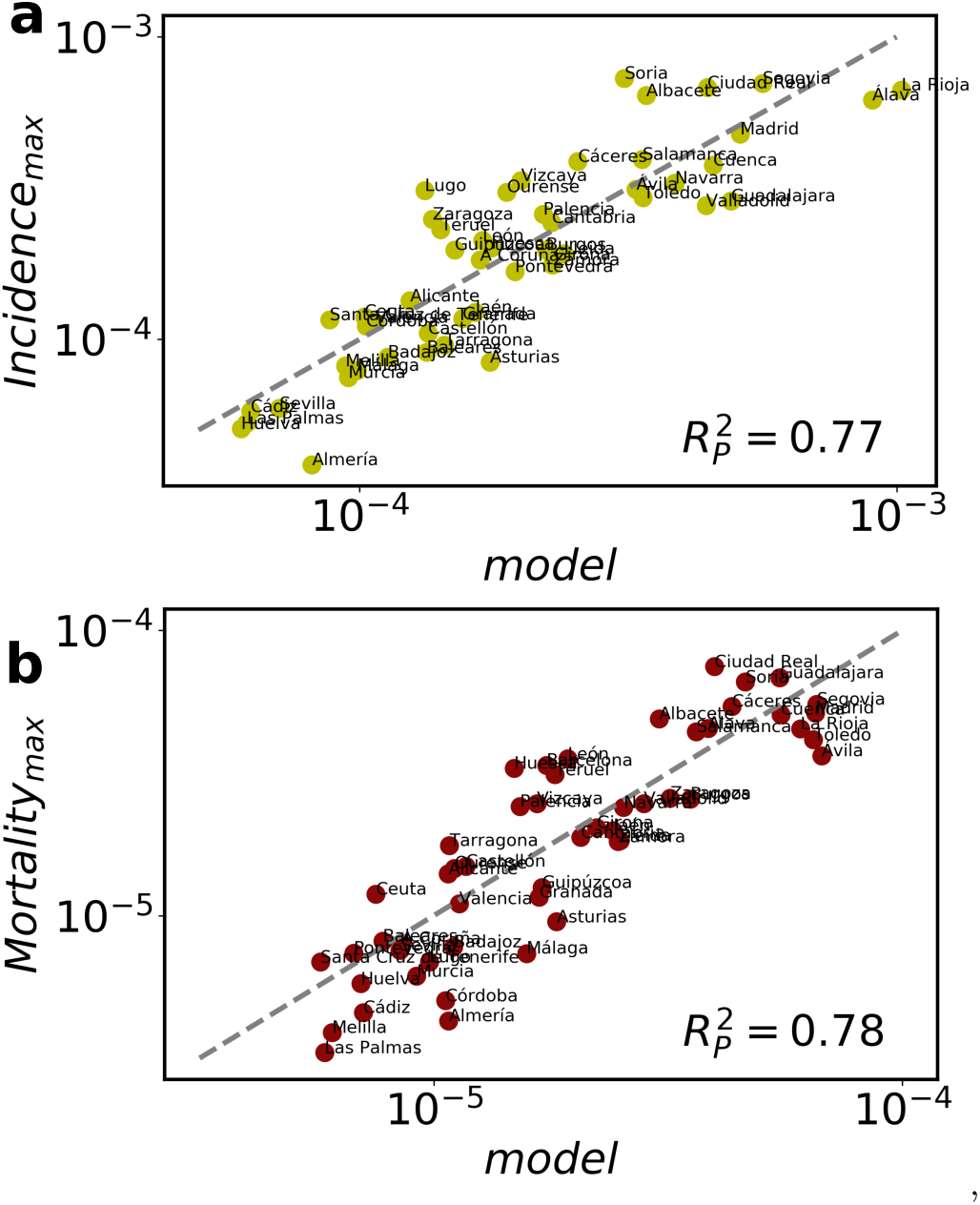
Statistical models. Comparison plots for the height of the a incidence and b mortality peaks. The x-axis displays the predictions of the corresponding statistical model and the y-axis the actual values in every province. The explained variance in Pearson coefficients is of 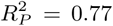 and 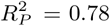, respectively.

**FIG. 7.**
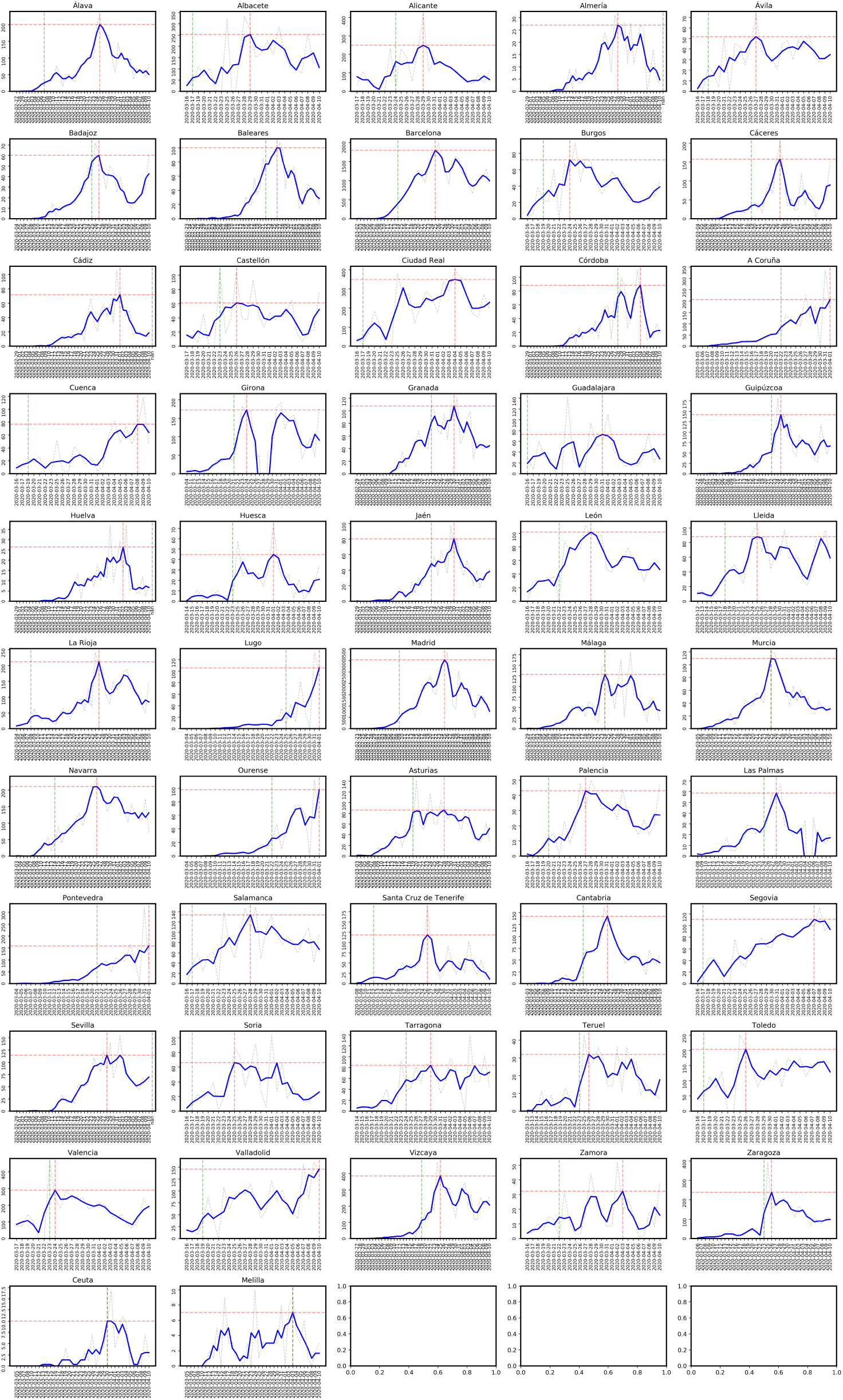
Epidemic curves of confirmed cases for the 52 provinces of Spain. Green dashed line to represent the incidence onset, red dashed line to represent the incidence peak time.

**FIG. 8.**
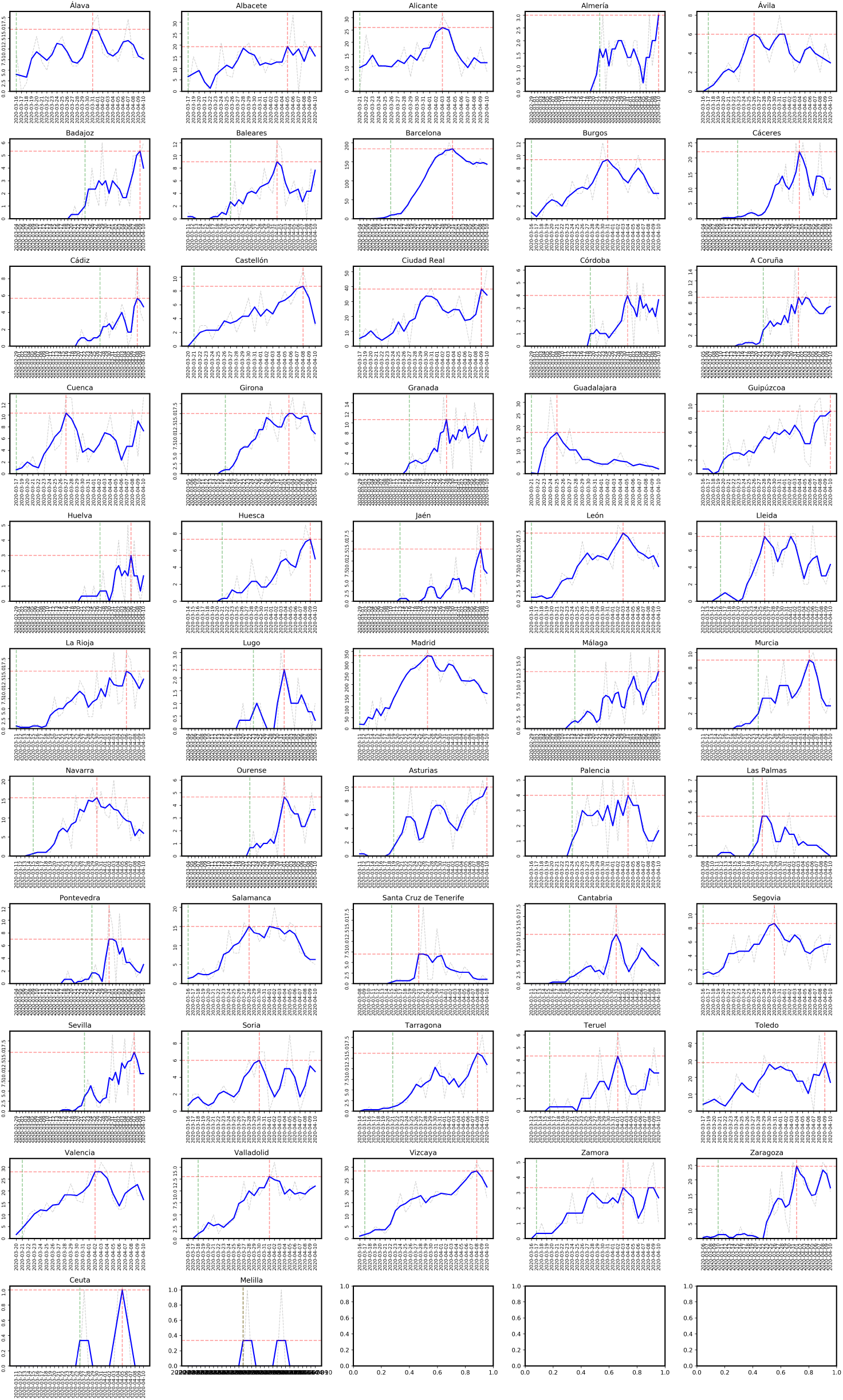
Epidemic curves of reported deaths for the 52 provinces of Spain. Green dashed line to represent the mortality onset, red dashed line to represent the mortality peak time.

**FIG. 9.**
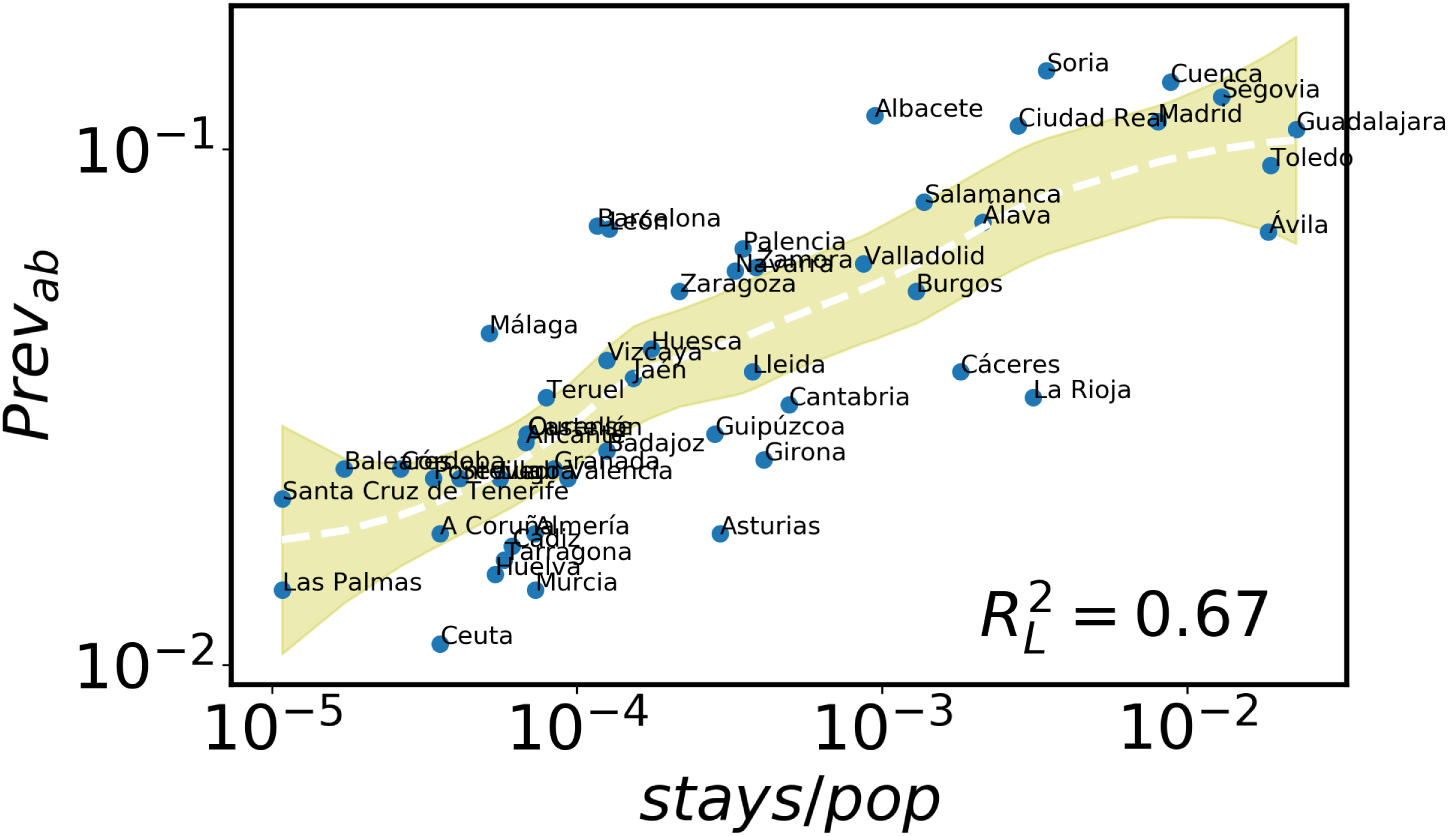
Stays per capita versus temporal size at May 13th 2020. Correlation plot of stays per capita of weekenders from Madrid to other provinces versus size of incidence in the local population.

**FIG. 10.**
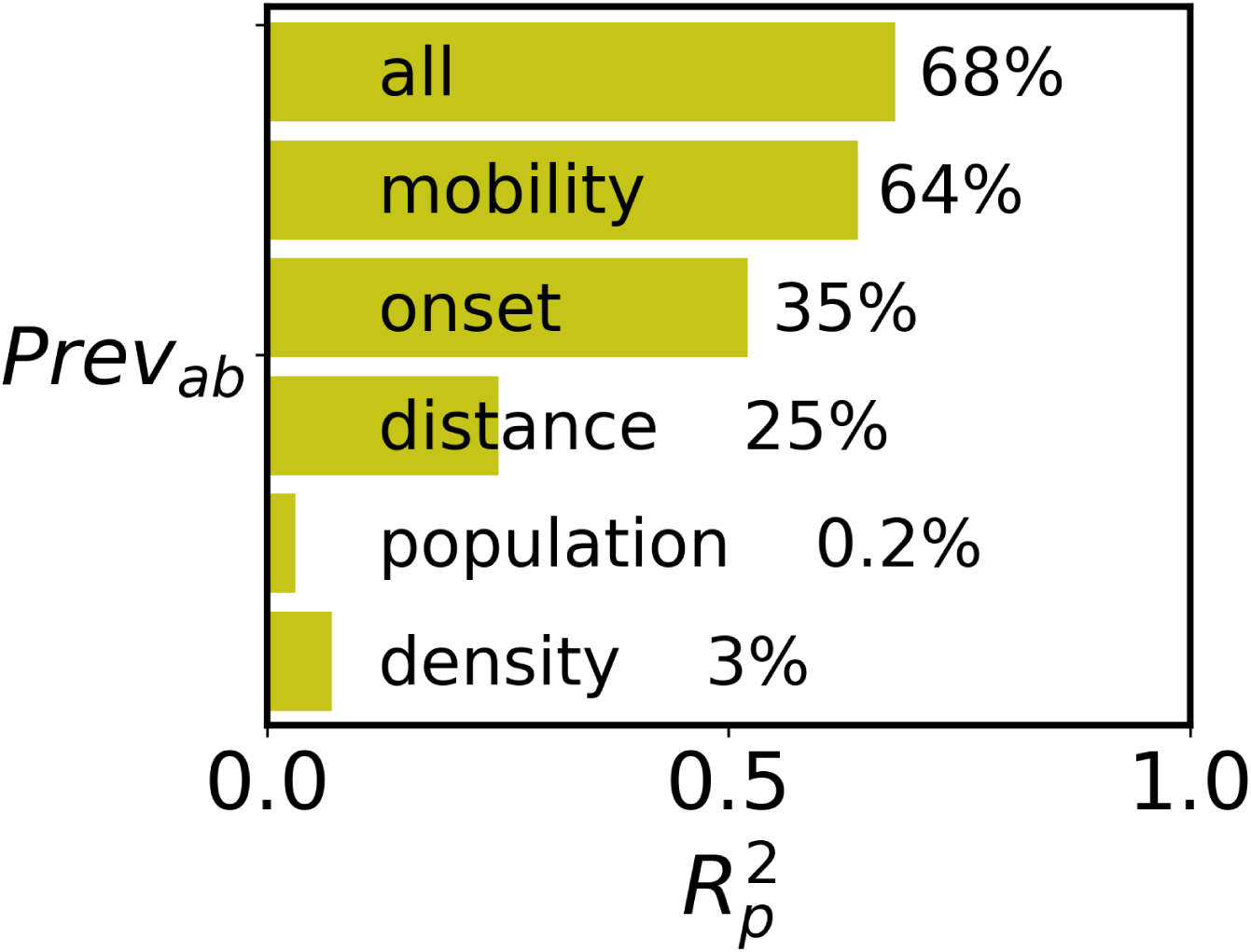
Correlation analysis of antibodies prevalence at May 13th 2020. Variance of local size at May 13th 2020 explained by mobility, population, population density and distance from Madrid of each province.

**FIG. 11.**
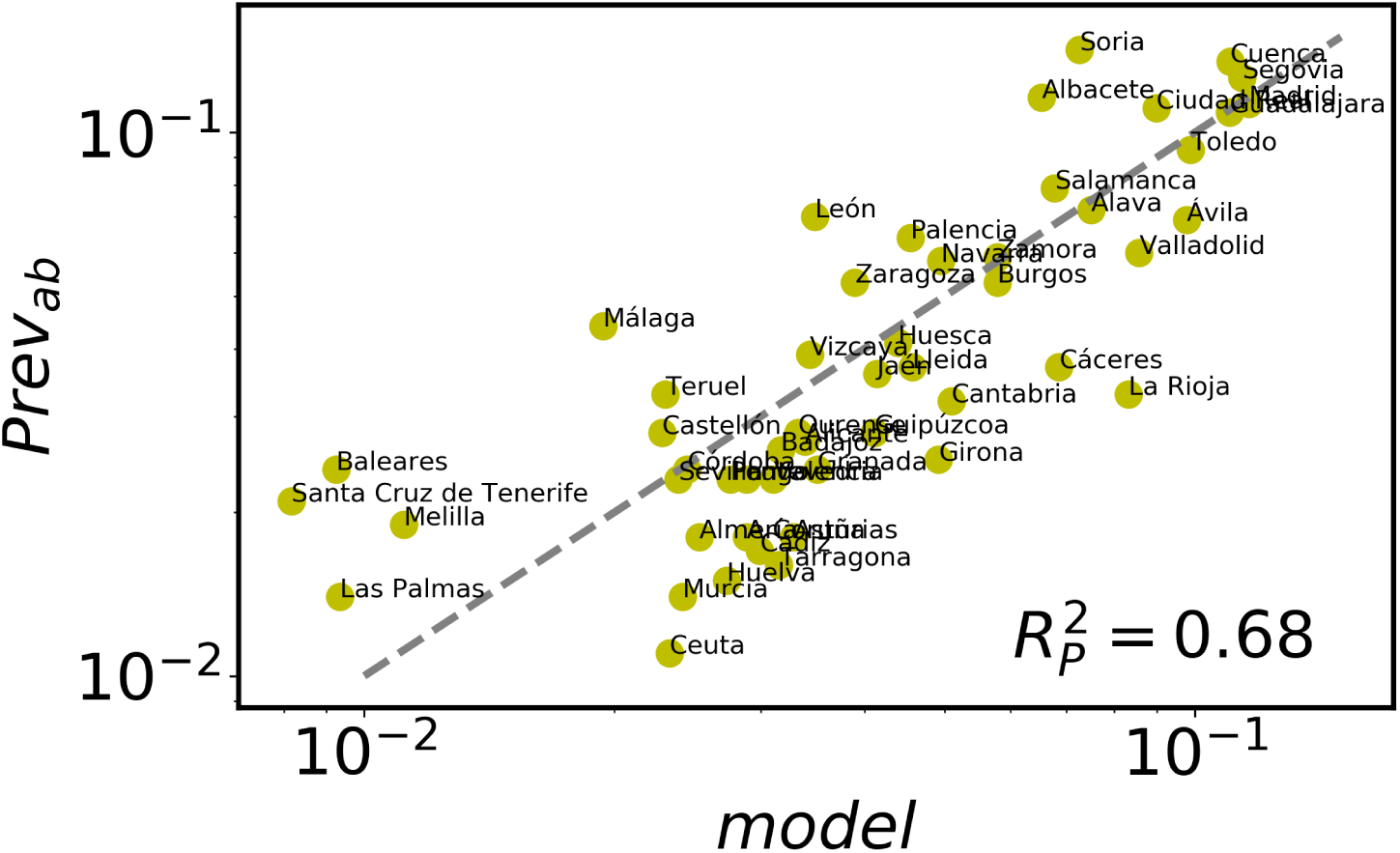
Multivariate model for the antibodies prevalence in each province.

Finally, the Spanish Ministry of Health has just released the first results of a study on the prevalence of antibody response in the population (https://www.mscbs.gob.es/gabinetePrensa/notaPrensa/pdf/13.05130520204528614.pdf). The analysis tested 60,983 individuals from different provinces, age groups and socio-economic contexts in a sample obtained with the same mechanisms as the census. The results are thus expected to be representative of the extension of the pandemia in the population. We have repeated the same analysis presented so far with this new data (see Supplementary Material, Figs. 9, 10, and 11 below). The mobility in terms of stays per capita explains 64% of the variance of the antibody prevalence (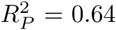), while the full multivariate model accounts for 68% and the comparison between the statistical model and the data yields 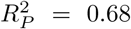. Despite the data of the prevalence analysis has been obtained much later than the peaks, these results strongly confirm the scenario we have found of dependence between the peaks (mortality and incidence) and the mobility.

## CONCLUSIONS

Understanding the impact of mobility on the early stages of spreading of an epidemic is key to design efficient public health responses. In this work, we address the question of how relevant is multi-seeding for the epidemic indicators in a population. The relation between a larger inflow of seeds and the epidemic size, heights of the incidence and mortality peaks is easy to hypothesize from a theoretical perspective. Is it, however, a reality? To test it, we perform a simple correlation analysis in the context of the present COVID-19 pandemic in Spain. The first local outbreak was declared in Madrid, attaining 100 confirmed cases on March 6, 2020. From then on, cases were detected in all the 52 provinces and autonomous cities of the country. The confinement measures were taken in a uniform way across the country with a first lockdown on Friday March 14, 2020, a stricter lockdown on March 29. The incidence peaks for most of the provinces occurred after the first lockdown and before the second. The fact that the lockdowns are uniform allows us to perform the analysis on multi-seeding in equal conditions. We find a clear relation between the mobility from/to Madrid and the heights of the peaks of incidence and mortality. Furthermore, the kind of mobility that better correlates is the one exhibited by weekenders one week before the local onset and, typically, before the confinement measures. The results are further confirmed by obtaining similar correlations between the prevalence of antibodies in the population measured in an independent study and the mobility. This supports the policies intended to target non-essential trips first and, only later, commuting flows (weekday travelers).

It is not surprising that a larger mobility to and from the area with the first outbreak accelerates the local onsets. But if the flow of trips per capita are high and frequent, the probability of importing more cases increases. Multi-seeds impact in different zones of the local social/contact network. The protection that bottlenecks in such contact network structure could bring is, thus, deactivated, which leads in turn to an enhancement of the contagions and a larger epidemic size with respect to a single or few seeds scenario. More cases imply larger mortality. These are theoretical arguments of the effect of multi-seeding. The main contribution of our work is to offer empirical evidence of such connections between multi-seeding and the severity of the epidemic at the population level. We hope that this study will highlight the importance of mobility in an epidemic situation, which goes beyond a first direct relation between arrival times and inflow of trips, and help stakeholders and decisionmakers to design more efficient responses.

## Data Availability

The information on mobility are available upon request with the permission of Kido Dynamics SA. The data on incidence and mortality can be downloaded from [29] and the one on the antibody prevalence from https://www.mscbs.gob.es/gabinetePrensa/notaPrensa/pdf/13.05130520204528614.pdf.

## DATA AVAILABILITY STATEMENT

The information on mobility are available upon request with the permission of Kido Dynamics SA. The data on incidence and mortality can be downloaded from [29] and the one on the antibody prevalence from https://www.mscbs.gob.es/gabinetePrensa/notaPrensa/pdf/13.0513

## ACKNOWLEDGEMENTS

M.M. is funded by the Conselleria d’Innovacio, Recerca i Turisme of the Government of the Balearic Islands and the European Social Fund with grant code FPI/2090/2018. M.M., S.M. and J.J.R. also acknowledge funding from the project Distancia-COVID (CSIC-COVID-19) of the CSIC funded by a contribution of AENA, from the Spanish Ministry of Science, Innovation and Universities, the AEI and FEDER (EU) under the grant PACSS (RTI2018-093732-B-C22) and the Maria de Maeztu program for Units of Excellence in R&D (MDM-2017-0711).

## References

[1] WHO COVID-19 Timeline. https://www.who.int/news-room/detail/27-04-2020-who-timelinecovid-19

[2] WHO COVID-19 Dashboard. https://covid19.who.int/

[3] Epicentre. COVID-19 Epi Dashboard. https://reports.msf.net/public/covid19/.

[4] https://www.lamoncloa.gob.es/consejodeministros/resumenes/Paginas/2020/14032020_alarma.aspx

[5] Hufnagel L, Brockmann D, Geisel T. Forecast and control of epidemics in a globalized world. Proc Natl Acad Sci USA. 2004; 101:15124 -15129. http://dx.doi.org/10.1073/pnas.0308344101

[6] Viboud C, Miller MA, Grenfell BT, Bjornstad ON, Si-monsen L. Air travel and the spread of influenza: important caveats. PLoS Med. 2006; 3: e503.

[7] Cooper BS, Pitman RJ, Edmunds WJ, Gay N. Delaying the international spread of pandemic influenza. PloS Med. 2006; 3: e12.

[8] Hollingsworth TD, Ferguson NM, Anderson RM. Will travel restrictions control the international spread of pandemic influenza? Nature Med. 2006; 12: 497–499.

[9] Ferguson NM et al. Strategies for mitigating an influenza pandemic. Nature 2006; 442: 448–452.

[10] Epstein JM et al. Controlling pandemic flu: the value of international air travel restrictions. PLoS ONE 2007; 2: e401.

[11] Balcan D, et al. Seasonal transmission potential and activity peaks of the new influenza A (H1N1): a Monte Carlo likelihood analysis based on human mobility. BMC Medicine. 2009;7:45

[12] P. Bajardi, et al. Human mobility networks, travel restrictions, and the global spread of 2009 H1N1 pandemic. PLoS ONE. 2011;6:e16591. http://dx.doi.org/10.1371/journal.pone.0016591

[13] M. Tizzoni, et al. Real-time numerical forecast of global epidemic spreading: casestudy of 2009 A/H1N1pdm. BMC Medicine. 2012;10:165

[14] S. Riley, K. Eames, V. Isham, D. Mollison, P. Trapman. Five challenges for spatial epidemic models. Epidemics. 2015; 10:68–71. doi.org/10.1016/j.epidem.2014.07.001.

[15] M.U.G. Kraemer, et al. The effect of human mobility and control measures on the COVID-19 epidemic in China. Science. 2020; 368:493–497. DOI: 10.1126/sci-ence.abb4218

[16] B.F. Maier, Dirk Brockmann. Effective containment explains subexponential growth in recent confirmed COVID-19 cases in China. Science. 2020;386:742–746. DOI: 10.1126/science.abb4557

[17] M. Chinazzi et al. The effect of travel restrictions on the spread of the 2019 novel coronavirus (COVID-19) outbreak. Science. 2020; 368: 395-400.

[18] A. Aleta, Y. Moreno. Evaluation of the potential incidence of COVID-19 and effectiveness of contention measures in Spain: a data-driven approach. medRxiv (2020). https://doi.org/10.1101/2020.03.01.20029801

[19] A. Arenas et al. A mathematical model for the spatiotemporal epidemic spreading of COVID19. (2020) https://doi.org/10.1101/2020.03.21.20040022

[20] B. Klein et al. Assessing changes in commuting and individual mobility in major metropolitan areas in the United States during the COVID-19 outbreak. (2020) https://www.m0bs-lab.0rg/upl0ads/6/7/8/7/6787877/assessing_mobility_changes_in_the_united_states_during_the_covid_19_outbreak.pdf

[21] M. Gilbert et al. ’’Preparedness and vulnerability of African countries against importations of COVID-19: a modelling study.” The Lancet. 2020;395: 871–877.

[22] T. Hasegawa, K. Nemoto. Outbreaks in susceptible-infected-removed epidemics with multiple seeds. Phys. Rev. E. 2016; 93: 032324.

[23] T. Hasegawa, K. Nemoto. Sudden spreading of infections in an epidemic model with a finite seed fraction. Eur. Phys. J. B. 2018; 91: 58. https://doi.org/10.1140/epjb/e2018-80343-3

[24] L. Wang L, J.T. Wu. Characterizing the dynamics underlying global spread of epidemics. Nat Commun. 2018; 15:218. DOI: 10.1038/s41467-017-02344-z.

[25] T. Sakai, et al. Geographic and temporal trends in influenza like illness, Japan, 1992-1999. Emerg. Infect. Dis. 2004; 10: 1822-1826.

[26] P. Coletti, et al. Shifting patterns of seasonal influenza epidemics. Sci Rep. 2018; 8: 12786. DOI: 10.1038/s41598-018-30949-x

[27] G. Pullano, et al. Novel coronavirus (2019-nCoV) early-stage importation risk to Europe, January 2020. Euro Surveill. 2020; 25: 2000057 DOI:10.2807/1560-7917.ES.2020.25.4.2000057.

[28] C. Dwork and A. Roth. The Algorithmic Foundations of Differential Privacy. Foundations and Trends in Theoretical Computer Science. 2014; 9: 211–407. DOI: 10.1561/0400000042.

[29] The data on cases and casualties have been obtained from the collaborative repository Es- COVID19data (https://github.com/montera34/escovid19data), which has collected information from: ‘analisi.transparenciacatalunya.cat’, ‘analisis.datosabiertos.jcyl.es’, ‘app.flourish.studio’,’ aragonhoy.aragon.es’,’aragonhoy.net’,’cadenaser.com’, ‘coronavirus.san.gva.es’,’galiciancovid19. info’,’govern.cat’,’infocamp.cat’,’public. flourish.studio’,’reusdigital.cat’,’sanidad. castillalamancha.es’,’tarragonadigital.com’, ‘www.acn.cat’, ‘www.aragonhoy.net’, ‘www.castillalamancha.es’, ‘www.diaridegirona.cat’, ‘www.diaridetarragona.com’, ‘www.diarimes.com’, ‘www.diariodelaltoaragon.es’, ‘www.diariodeteruel.es’, ‘www.elperiodicoextremadura.com’, ‘www.euskadi.eus’, ‘www.gva.es’, ‘www.heraldo.es’, ‘www.hoy.es’, ‘www.juntadeandalucia.es’, ‘www.juntaex.es’, ‘www.lacomarca.net’, ‘www.naciodigital.cat’, ‘www.radiobalaguer.cat’, ‘www.san.gva.es’, ‘www.segre.com’, ‘www3.gobiernodecanarias.org’

[30] https://elpais.com/sociedad/2020-03-06/mas-de-60-personas-se-contagiaron-a-la-vez-en-un-funeral-en-vitoria.html

